# Identifying At-Risk Communities and Key Vulnerability Indicators in the COVID-19 Pandemic

**DOI:** 10.1101/2021.09.19.21263805

**Authors:** Savannah Thais, Shaine Leibowitz, Alejandra Rios Gutierrez, Alexandra Passarelli, Stephanie Santo, Nora Shipp

## Abstract

Throughout the COVID-19 pandemic, certain communities have been disproportionately exposed to detrimental health outcomes and socioeconomic injuries. Quantifying community needs is crucial for identifying testing and service deserts, effectively allocating resources, and informing funding and decision making. We have constructed research-driven metrics measuring the public health and economic impacts of COVID-19 on vulnerable populations. In this work we further examine and validate these indices by training supervised models to predict proxy outcomes and analyzing the feature importances to identify gaps in our original metric design. The indices analyzed in this work are unique among COVID-19 risk assessments due to their robust integration of disparate data sources. Together, they enable more effective responses to COVID-19 driven health inequities.

## 1 Introduction

The COVID-19 pandemic has resulted in devastating global health, economic, and social impacts. Research has shown that in the United States this virus affects certain communities more severely: minority communities are experiencing higher death rates [Wiemers *et al*., 2020], low-income communities are facing housing crises and food shortages [Raifman and Raifman, 2020], and rural communities suffer from limited access to healthcare resources [Douthit *et al*., 2015]. These disparities have grown larger as the pandemic continues, and, despite the discovery of an effective vaccine, they will have long-lasting consequences for the affected communities.

Prior to COVID-19, the US Center for Disease Control’s (CDC) Social Vulnerability Index (SVI) [Flanagan *et al*., 2018] was commonly employed to analyze community data for policy and planning in governmental and community public health initiatives. To incorporate the impact of the pandemic, the SVI has since been integrated with COVID-19 case and death rate data [Khazanchi *et al*., 2020] and other work has explored creating new vulnerability estimates specifically for estimating COVID-19 effects [Baer *et al*., 2020]. The work presented in this paper surpasses previous work by developing a new suite of metrics to quantify different models of vulnerability to disparate COVID-19 impacts, rather than focusing on general vulnerability to negative health outcomes, and through extensive validation, analysis, and interpretation of the metrics.

## 2 The COVID-19 Community Vulnerability Metrics

The Community Vulnerability Index (CVI) aggregates county-level data in the US for three distinct metrics: COVID-19 case severity (Severity), risk of economic harm (Economic Harm), and need for mobile health resources (Mobile Health). Each metric is a weighted combination of quantile-normalized variables. The construction of each metric, including which variables to incorporate and how to weigh them relative to each other, is informed by an extensive review of public health, social science, and urban planning literature. This project is entirely open source and thus subject to the constraints of publicly available data; in some cases the metric went through several iterations until a suitable proxy was found for the initial variable.

The Severity metric measures the risk of hospitalization as a benchmark for severe COVID-19 complications in a county. While the number of COVID-19 hospitalizations would have been the preferred base indicator for this metric, a nationwide data set on hospitalizations was unavailable. Therefore, the base indicator is the number of COVID-19 cases (‘Covid Cases’) which was assigned the lowest weight of 1, in recognition of the indicator’s limitations. Given the nationwide testing shortage at the beginning of the pandemic and the asymptomatic nature of COVID-19, case numbers do not directly correlate to hospitalization numbers [Gao *et al*., 2021].

Accompanying this base indicator are comorbidities found to be prevalent and associated with general COVID-19 hospitalizations as opposed to strictly intensive care unit (ICU) admission, in-hospital death, or invasive mechanical ventilation. The selections were made according to pre-print and peer-reviewed articles from the early stages of the pandemic from March 2020 to June 2020. Diabetes, obesity, and cardiovascular disease are highly prevalent among and strongly associated with COVID-19 hospital admission, ICU admission, and in-hospital death [Lodigiani *et al*., 2020] [Al-Sabah *et al*., 2020][Yang *et al*., 2020]. Interestingly, hypertension is both highly prevalent among COVID-19 hospital admission and also protective against ICU admission and in-hospital death for COVID-19 [Richardson *et al*., 2020]. Therefore, ‘% Diagnosed Diabetes’, ‘% Adults with Obesity’, ‘Heart Disease Death Rate’, and ‘Hypertension Death Rate’ share the highest weight of 4 in the metric. While diabetes and obesity are cyclical pathologies, peer-reviewed and pre-print papers reported both diseases among severely ill COVID-19 patients. [Al-Sabah *et al*., 2020] [Hajifathalian *et al*., 2020].

Contrary to early suspicions that pre-existing lung issues would exacerbate COVID-19 symptoms [Collaborative *et al*., 2020], many respiratory conditions did not have strong estimated associations nor high frequencies among COVID-19 hospital-admitted patients. For example, asthma was not found to have any statistically significant association with any COVID-19 severity outcomes. Notably, Chronic Obstructive Pulmonary Disease (COPD) was associated with COVID-19 outcomes, but not as strongly as previously mentioned metric indicators, and consequently was given a lower weight of 3. ‘% Smokers’ presented the weakest association with COVID-19 hospitalization, and was given the lowest weight of 1 [Collaborative *et al*., 2020].

Finally, ‘% Adults 65 and Older’ in a county is an effect modifier for all indicators in the metric. Elderly age increased the frequency and association of comorbidities with severe COVID-19 outcomes. Therefore, ‘% Adults 65 and Older’ was given a weight of 4 [Zhou *et al*., 2020].

The Economic Harm metric measures a county’s risk of severe, negative economic impact due to COVID-19. The metric considers several traditional economic development indicators: poverty, income, educational attainment, and unemployment. The variables ‘% Below Poverty’ and ‘Median Household Income’ capture low-income communities’ pre-existing economic needs [Drobniak, 2012]. Measuring pre-existing need is essential for understanding which communities are less resilient and might take longer to recover from a recession. Moreover, ‘% No College Degree’ was chosen as a measure for educational attainment because college graduates tend to have higher job security that allows for telecommuting. Unemployment is split into two indicators that aim to capture a more complex understanding of job losses amid the pandemic [Mikolai *et al*., 2020]. The ‘Unemployment Rate’ is inclusive of permanent job losers, job leavers, and people that are temporarily laid-off. ‘% Not in Labor Force’, which consists of marginally attached and discouraged workers, aims to capture people that are not counted in traditional unemployment measures since they have discontinued their job search or have chosen to stay out of the workforce during the pandemic [Bauer *et al*., 2020] [of Labor Statistics, 2020]. Additionally, the metric takes into account people with precarious jobs, defined as non-standard or temporary employment, by including ‘% Part-time’ and ‘% Self-Employed’ [Bartik *et al*., 2020a]. Ideally, shift, temporary, gig, and seasonal workers would also be included, but nationwide datasets for these worker types were unavailable.

Finally, the metric considers place-based characteristics of a county. The ‘% Jobs in Leisure and Hospitality’ captures the job outlook for the hardest-hit industry during the pandemic [Forsythe *et al*., 2020]. This industry continues to struggle the most with recovery, and counties dependent on it remain the most devastated [Muro *et al*., 2020]. For this metric, all variables have equal weight as this work is one of the earliest comprehensive studies of COVID-19’s community economic impact, and additional analysis, including work described in Section 3.3, is needed to understand relative feature importance.

The Mobile Health metric measures the community need for non-traditional healthcare delivery services at the county level. The literature review for the metric was inclusive of mobile health clinics (physical clinics on wheels), telehealth services (virtual health services that connect patients to care), and health app solutions (technology that allows healthcare personnel to monitor patients’ symptoms remotely).

A critical category of indicators in the metric describes different measurements for physical isolation from healthcare services. The indicators ‘Primary Care Physicians Rate’ and ‘Number of Hospitals’ were given a weight of -3 to highlight the lack of healthcare infrastructure in a county [Yu *et al*., 2017]. Very rural communities are inherently isolated, including isolation from healthcare infrastructure, and, therefore, ‘% Rural’ was also given a weight of 3 [Malone *et al*., 2020]. The metric also considers the availability of transportation to connect to traditional healthcare services by including ‘% Households Without a Car’ with a weight of 2 and ‘% Workers Commuting by Public Transit’ with a weight of - 2 [Yu *et al*., 2017]. The latter two indicators weigh lower than the physical barriers to healthcare because access to transportation does not directly result in uptake of healthcare services, especially if services are few and far between.

Two intertwined barriers to healthcare are administrative and cultural barriers. Those without insurance, without strong English language skills, and ethnic minority groups are more hesitant to seek healthcare services, and navigate the US healthcare system at a disadvantage without linguistically and culturally appropriate care [Yu *et al*., 2017] [Malone *et al*., 2020].’% Without Health Insurance’ and ‘% Limited English Proficiency’ are given a weight of 2, whereas ‘% Non-white’ is given a weight of 1 since racial data is more distal proxy for place-based disenfranchisement [Price *et al*., 2013].

Certain vulnerable and under-served populations who traditionally lack access to healthcare are also considered in the metric: ‘% Veterans in Civilian Adult Population’, ‘% Adults 65 and Older’, ‘% People with Disabilities’, and the ‘Opioid Death Rate’ [Malone *et al*., 2020]. The selection of groups to include in this category was limited by dataset availability. Elders and people with disabilities were weighed more (2 instead of 1) because of their more proximate inability to physically reach healthcare systems [Chauhan *et al*., 2020]. Finally, the metric includes the general health status of a county: ‘% Fair or Poor Health’, with a weight of 1.

Accompanying the mobile health need metric are selectable overlays that visualize ‘% With Home Internet Access’ and ‘% With Smartphone/Tablets’. While the metric incorporates a need for various non-traditional healthcare services, the two variables measure internet access capabilities, which are only relevant for telehealth visits and mobile health apps [Kruse *et al*., 2018] [Carroll *et al*., 2017]. Additionally, keeping the two variables separate empowers data users to decide what type of service to deploy and to where.

The variables that have been described in this section will henceforth be referred to as the Initial Variables. Organized tables of these variables and their weights are included in the Appendix.

## 3 Supervised Learning for Proxy Outcomes

To inform the next iteration of CVI metrics, we implemented supervised learning models to predict proxy outcomes. We sought to assess our current feature weights, quantify the predictive power of the included variables, and discover any information gaps in the initial metric construction.

### 3.1 Methodology

We chose a proxy outcome for each metric to serve as the learned predicted output for a supervised learning algorithm. The proxy outcomes were selected based on relevant literature that supported the selected outcome as a strong indicator of our intended measurement. In order to evaluate our current features (i.e. the Initial Variables), we compared them to algorithmically selected feature set. Starting with the full CVI dataset, which includes all metric variables and additional Social Determinants of Health [the U.S. Department of Health *et al*., 2021], we narrowed our algorithmically selected feature set by selecting the most important features according to XGBoost’s F-score. We randomly split the dataset into 80% training and 20% test. The XGBoost model underwent hyper-parameter tuning with k=5 cross-validation. The XGBoost Most Important Features were tested for correlation and Predictive Power Score (PPS) where highly correlated variables were removed from the comparison set.

The Initial Variables and XGBoost Most Important Features were compared by training Multilayer Perceptrons (MLPs) to predict the proxy outcome and evaluated with Root Mean Squared Error (RMSE) on the test dataset. The training set was randomly split again into 80% training and 20% validation. A dummy baseline was calculated as the error on the average outcome of the training data. The final network architecture is as follows: dropout (0.2) on the input layer, 2 hidden layers, and 25 to 100 training epochs with early stopping due to validation dataset performance. To explain the output of the MLPs, we applied SHapley Additive exPlanations (SHAP) [Lundberg and Lee, 2017] and further examined the variables with the highest SHAP value magnitudes.

### 3.2 Severity

The proxy severity outcome is COVID-19 Hospitalization Rate per 100,000 population where the hospitalizations were measured cumulatively from March 2020 to April 2021. The distribution of the proxy outcome is featured in Figure 1. Due to data accessibility and accuracy constraints, we only obtained data on 5 states totaling 527 counties: Florida [the Florida Department of Health Open Data, 2020], Georgia [the Cobb County COVID-19 Resources, 2020], Tennessee [the Tennessee State Data Center and Research, 2020], Virginia [the Virginia Open Data Portal, 2020], and Wisconsin [the Wisconsin Department of Health Services, 2020]. This proxy outcome was the preferred base indicator for the Severity metric.

**Figure 1:**
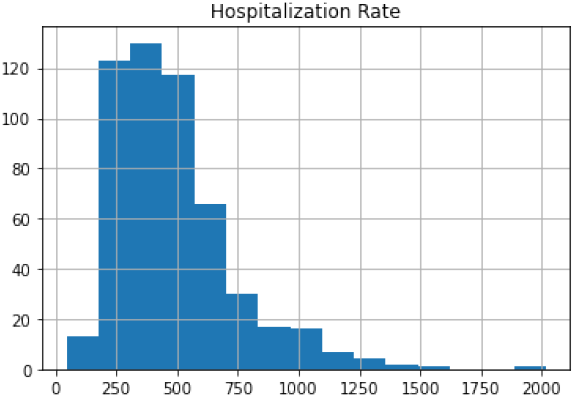
Distribution of Proxy Outcome (Severity)

The most important features according to XGBoost are shown in Table 1. ‘% Children Uninsured’ was removed from the comparison set due to its unsurprisingly high correlation with ‘% Without Health Insurance’.

**Table 1:**
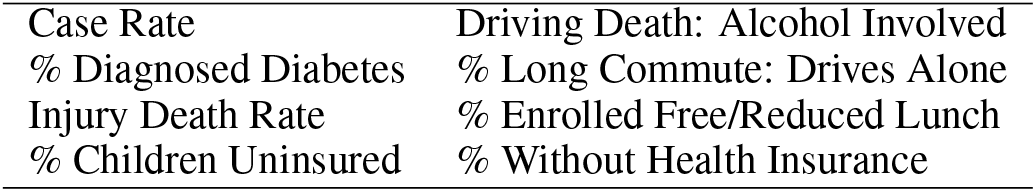
XGBoost Most Important Features (Severity)

|  |  |
| --- | --- |
| Case Rate | Driving Death: Alcohol Involved |
| % Diagnosed Diabetes | % Long Commute: Drives Alone |
| Injury Death Rate | % Enrolled Free/Reduced Lunch |
| % Children Uninsured | % Without Health Insurance |

As Table 2 exhibits, the XGBoost Most Important Features perform better than the Initial Variables. As Figure 2 exhibits, ‘% Enrolled in Free or Reduced Lunch’ is the variable that most explains the MLP’s output. As this feature can be considered as an indicator of poverty, we included other common indicators of poverty: ‘% Below Poverty’, ‘Unemployment Rate’, and ‘% Children in Poverty’ one at a time for comparison. However, ‘% Enrolled in Free or Reduced Lunch’ was substantially more predictive than other poverty indicators (see Section 4.1 for further discussion).

**Table 2:** RMSE on test dataset (Severity)

| Feature Set | RMSE |
| --- | --- |
| Dummy Baseline | 2358.6 |
| Initial Variables | 228.2 |
| + % Enrolled in Free or Reduced Lunch | 198.3 |
| + % Below Poverty | 221.7 |
| + Unemployment Rate | 231.9 |
| + % Children in Poverty | 229.6 |
| XGBoost Most Important Features | 183.1 |

**Figure 2:**
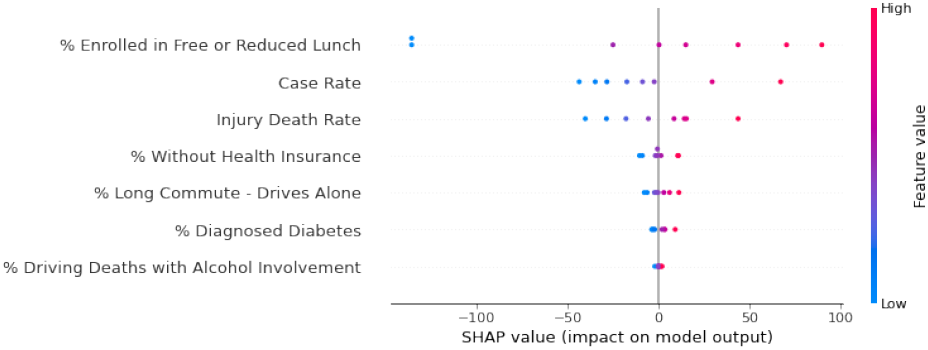
SHAP Values for MLP with the XGBoost Most Important Features as Input (Severity)

### 3.3 Economic Harm

As a measure of individual economic status, we use a proxy outcome of cumulative unemployment initial claims between January and November 2020 per 100 people in the 2019 labor force [Chetty *et al*., 2021]. The distribution of the proxy outcome is shown in Figure 3. The proxy differs from the pre-covid unemployment rates included in the original metric to estimate existing economic precarity in the county.

**Figure 3:**
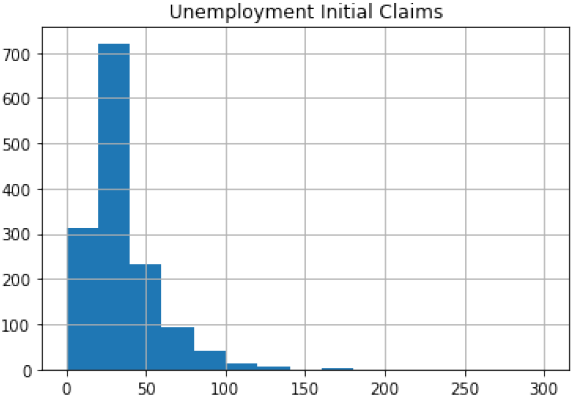
Distribution of Proxy Outcome (Economic Harm)

The most important features according to XGBoost are shown in Table 3. Again, ‘% Children Uninsured’ was removed from the comparison set due to its high correlation with ‘% Without Health Insurance’.

**Table 3:** XGBoost Most Important Features (Economic Harm)

|  |  |
| --- | --- |
| Average Daily PM2.5 | % Children Uninsured |
| Unemployment Rate | % American or Alaskan Native |
| % Insufficient Sleep | % Excessive Drinking |
| % Self-Employed | % Without Health Insurance |
|  | % Severe Housing Cost Burden |

As Table 4 exhibits, the XGBoost Most Important Features perform better than the Initial Variables. As shown in Figure 4, ‘% Self-Employed’ is the variable that most explained the MLP’s output. Since ‘% Self-Employed’ is already in the Initial Variables, we look at including the next most important feature ‘% Insufficient Sleep’. Adding this variable to the Initial Variables in another MLP has a slight but most likely insignificant effect on the error.

**Table 4:** RMSE on test dataset (Economic Harm)

| Feature Set | RMSE |
| --- | --- |
| Dummy Baseline | 338.7 |
| Initial Variables | 22.2 |
| + % Insufficient Sleep | 21.6 |
| XGBoost Most Important Features | 14.7 |

**Figure 4:**
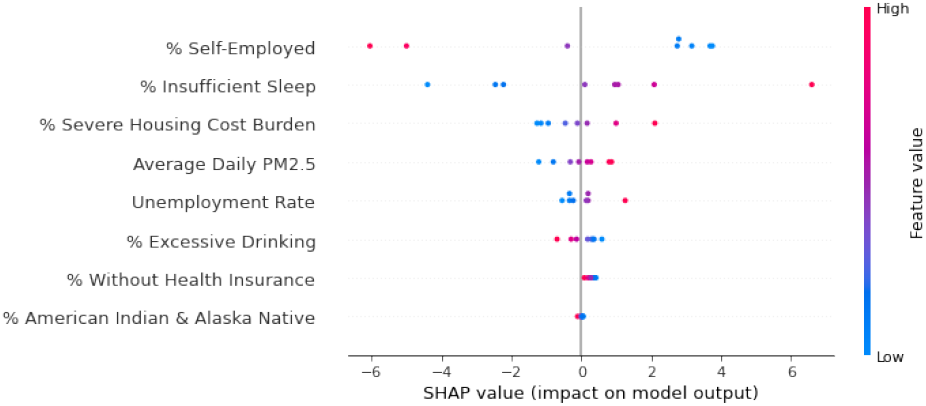
SHAP Values for MLP with the XGBoost Most Important Features as Input (Economic Harm)

### 3.4 Mobile Health

Counties with a low number of hospitals have a higher need for mobile healthcare services [Yu *et al*., 2017]; thus, the proxy mobile health outcome is Ratio of Hospitals per 100,000 population. The distribution is shown in Figure 5. Though ‘Number of Hospitals’ is one of the Initial Variables, it was removed from the feature sets for this analysis.

**Figure 5:**
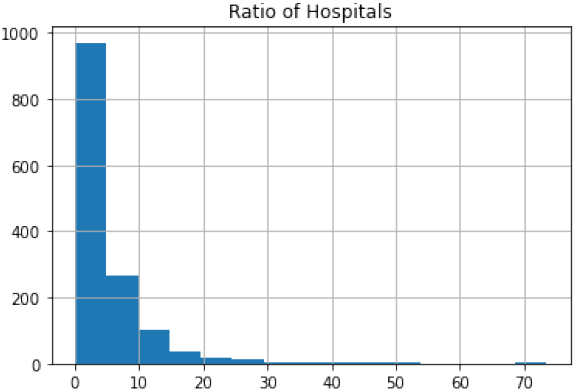
Distribution of Proxy Outcome (Mobile Health)

The most important features according to XGBoost are shown in Table 5. ‘Population’ and ‘% With Home Internet Access’ were both removed due to their high PPS with ‘Deaths’ (here, ‘Deaths’ only includes deaths attributed to COVID-19 measured daily). We expect ‘Deaths’ and ‘% With Home Internet Access’ to be correlated with ‘Population’ because ‘Deaths’ is a count rather than a rate and because more densely populated (non-rural) regions tend to have higher rates of internet access availability and adoption [Whitacre, 2010].

**Table 5:** XGBoost Most Important Features (Mobile Health)

|  |  |
| --- | --- |
| Population | Primary Care Physicians Rate |
| Deaths | % With Access to Exercise Opp. |
| % Vaccinated | % With Home Internet Access |
|  | % Long Commute - Drives Alone |
|  | Other Primary Care Provider Ratio |
|  | Social Association Rate |

As Table 6 exhibits, the XGBoost Most Important Features perform better than the Initial Variables. As can be seen in Figure 6, ‘Deaths’ is the variable that most explained the MLP’s outputs. Adding this variable to the Initial Variables decreases the error to be on par with the XGBoost Most Important Features.

**Table 6:** RMSE on test dataset (Mobile Health)

| Feature Set | RMSE |
| --- | --- |
| Dummy Baseline | 106.9 |
| Initial Variables | 5.0 |
| + Deaths | 4.3 |
| XGBoost Most Important Features | 4.3 |

**Figure 6:**
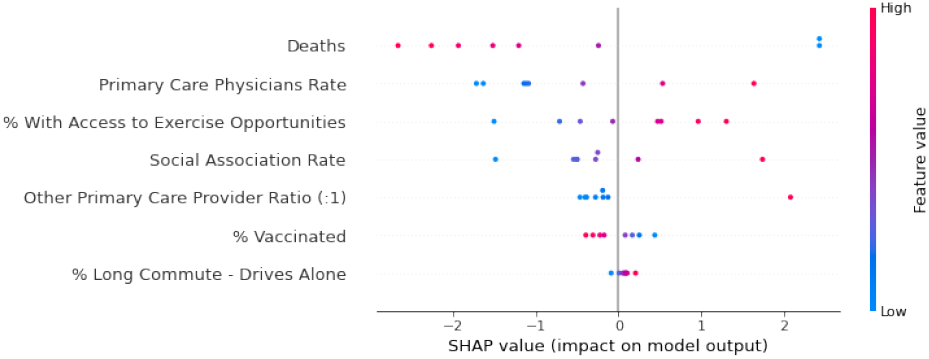
SHAP Values for MLP with the XGBoost Most Important Features as Input (Mobile Health)

## 4 Discussion

There are several important takeaways from this study. Understanding the relative feature importances in the supervised models can help fine-tune the weights of the vulnerability metrics, particularly in the case of the Economic Harm metric where relevant literature was unavailable. Additionally, comparing the XGBoost important feature sets to the original vulnerability metrics can expose information gaps and improve the precision of our metrics. However, it is well known that many Social Determinants of Health (SDHs) are strongly correlated or are downstream effects of the root health or policy causes. Furthermore, uncertainty is inherent in the selection of proxy outcomes that do not always capture the entire intended goal of the vulnerability metric. Thus, it is critical that we do not naively add any high-scoring feature to the vulnerability model and expect improved accuracy. In the following section we discuss key results of the study and explore possible causal pathways and public health implications of identified informative variables.

### 4.1 Severity Metric

The proxy outcome for the Severity metric analysis (hospitalizations during COVID) is the closest to the intended application of the metric. Thus, we carefully consider the XG-Boost identified important features for inclusion in the metric. However, we used a limited dataset for this initial model with only 5 states, all located in the South or Midwest. Public health policy and community health measures vary widely across geographic regions; thus we plan to seek additional data sources and scale up this study before making final adjustments to the Severity metric.

Only two of the original metric features, ‘Case Rate’ and ‘% Diagnosed Diabetes’, were identified by XGBoost. ‘Case Rate’ is the most powerful predictor of severe COVID-19 cases in the supervised model, suggesting the prevalence of COVID-19 in an area is more important than the prevalence of pre-existing comorbidities; we can consider increasing the relative weight of ‘Case Rate’ in future iterations of the Severity metric. Interestingly, only one of the well-studied COVID-19 comorbidities, ‘% Diagnosed Diabetes’ is identified by XGBoost. However, ‘% Without Health Insurance’ was identified and studies have demonstrated an association between lack of insurance and increased mortality or disease severity due to the other prominent COVID-19 comorbidities: heart disease, hypertension, and COPD [Brooks *et al*., 2010][Song *et al*., 2020]. We thus conclude that including ‘% Without Health Insurance’ in the Severity metric captures additional relevant information: disease prevalence combined with access to healthcare are indicative of severe disease and mortality.

The most important feature identified by XGBoost was ‘% Enrolled in Free and Reduced Lunch’. Initially, we postulated that this served as a measurement of poverty, as higher rates of COVID-19 deaths are associated with poorer counties [Finch and Hernández Finch, 2020]. However, including other traditional measurements of poverty did not reduce the RMSE (see Table 2). In particular, including ‘% Children in Poverty’ did not reduce the RMSE despite focusing on the child recipients of Free and Reduced Lunch programs. In addition to being an indicator of poverty, Free and Reduced Lunch programs are an effort to mitigate childhood food insecurity, which is often indicative of family-level food insecurity [Gundersen *et al*., 2012]. Amongst adults, food insecurity is associated with increased rates of and complications due to chronic diseases (including many comorbidities of COVID-19) [Seligman *et al*., 2010]. Furthermore, food insecurity is not always associated with living below the poverty line and can have additional infrastructure and food access causes [Wight *et al*., 2014]. We thus conclude that the variable ‘% Enrolled in Free and Reduced Lunch’ captures important information related to the causes and severity of key COVID-19 comorbidities and other relevant health impacts and will include it in future iterations of the Severity metric.

### 4.2 Economic Harm Metric

The proxy outcome for the Economic Harm metric analysis (unemployment initial claims rates during COVID) is an excellent proxy for the individual level economic impact of COVID-19. Only two of the original metric features: ‘Unemployment Rate’ and ‘% Self Employed’ where identified by XGBoost. Both have impactful SHAP scores, providing initial indication that they should be highly weighted in the Economic Harm metric. Interestingly though, ‘% Self Employed’ is inversely correlated with the proxy outcome. We postulate that this is because although self employed workers were able to file for unemployment after the CARES Act was passed in March 2020, many who continued to work but with reduced hours were instead eligible for a different type of aid: Self-Employment Income Support Scheme grants which is not captured by the proxy outcome variable. This demonstrates a shortcoming of our selected proxy variable as it is well documented that self employed individuals were disproportionately impacted by COVID-19 [Bartik *et al*., 2020b].

Other impactful XGBoost identified variables include ‘% Insufficient Sleep’, ‘% Severe Housing Cost Burden’, and ‘Average Daily PM2.5’. ‘% Severe Housing Cost Burden’ (the percentage of households in a county paying more than 50% of their income on housing makes sense as a possible causal predictor of economic harm. Households with severe housing cost burdens are more likely to forgo healthcare and are less likely to have savings or emergency funds, making them more vulnerable to the economic and health impacts of the COVID-19 pandemic [Trusts, 2018]. We will consider including ‘% Severe Housing Cost Burden’ in future iterations of the Economic Harm metric. Insufficient sleep has been previously studied as an economic indicator due to the impact lack of sleep has on school and labor market success and public health; in fact, one study estimates that insufficient sleep amongst the US working population costs the economy up to $411 billion per year [Hafner *et al*., 2016] due to decreased productivity and missed workdays. However, additional analysis of how sleep behavior has changed during the COVID-19 pandemic and the impact of insufficient sleep on local economies is necessary to decide if ‘% Insufficient Sleep’ should be included in the Economic Harm metric; notably, including it with the original metric features did not substantially reduce the RMSE of the supervised model (see Table 4). Finally, although air pollution, specifically the average daily density of fine particulate matter (‘Average Daily PM2.5), has been causally connected to decreased lung function and adverse pulmonary effects, and is known to increase premature death risk [Pope *et al*., 2008], we are unable to find a possible causal link between this variable and local economic impact. It will not be included in future iterations of the Economic Harm metric.

### 4.3 Mobile Health Need Metric

The proxy outcome for the Mobile Health Needs metric analysis (hospitals per 100,000 population) is the farthest removed from the intended goal of the original metric as it does not capture information about hospital accessibility or other forms of health coverage. In fact, it is in many ways the inverse of what the Mobile Health Needs metric seeks to capture as it describes a presence, rather than lack, of healthcare availability. Nonetheless, this study can provide insights into fine-tuning the Mobile Health Needs metric by examining the inverse of the SHAP values calculated from the supervised model. Only two of the original metric features, ‘Primary Care Physicians Rate’ and ‘% With Home Internet Access’, were identified by XGBoost (although currently ‘% With Home Internet Access’ is included as an overlay rather than directly incorporated into the metric). ‘Primary Care Physicians Rate’ has a large SHAP value, supporting it being one of the highest (negatively) weighted variables in the original metric. ‘Other Primary Care Provider Ratio’, which describes access to non-physician-based care, such as nurse practitioners or physician assistants, was also identified by XGBoost and has a large SHAP value. We plan to incorporate this variable into future versions of the metric but require additional study on the interplay with ‘Primary Care Physicians Rate’ and poverty indicators as a majority of non-physician-based care in an area can still be indicative of shortcomings of the local healthcare infrastructure.

‘Deaths’ (daily COVID-19 death counts) was the XGBoost identified feature with the largest SHAP value. The SHAP value is negatively correlated with the proxy outcome, which intuitively makes sense as areas with fewer healthcare resources have higher rates of poor health [Riley, 2012] and were more likely to overwhelm existing healthcare infrastructure during a COVID outbreak [Miller *et al*., 2020]. However, the COVID-19 death rate in different counties is also highly dependent on public health policy implementation and adherence, so we will not include ‘Deaths’ in future versions of the Mobile Health Needs metric. Interestingly, the ‘% Vaccinated’ SHAP value is also inversely correlated with the proxy outcome. This is possibly due to many COVID-19 vaccination clinics being setup in pharmacies, community centers, and other other ‘pop-up’ locations, rather than solely in hospitals, however this requires further study. ‘Access to Exercise Opportunities’ (percentage of population with adequate access to locations for physical activity including sidewalks, parks, and gyms) and ‘Social Association Rate’ (number of membership associations per 10,000 population) also have large SHAP values. Both variables describe access to social and physical infrastructure that enable healthy behavior and improved health outcomes [House, 2001][Jones *et al*., 2015]. It is possible that a lack of these resources also indicated a need for additional mobile healthcare resources, in which case including these variables would improve our Mobile Health metric by augmenting the more traditional variables of ‘Number of Hospitals’ and ‘Primary Care Physicians Rate’. However, additional literature review on the background of these variables and their causal impact on community well-being is needed.

### 4.4 Implications for Community Vulnerability Assessments

SDHs are often considered collectively to assess a community’s overall health and risk of adverse effects. Our work demonstrates that carefully constructed subsets can accurately quantify specific risk and need types. This work also highlights the importance of on-going exchange between statistical analyses and domain knowledge. There was not complete overlap between our hand-selected feature sets and the XGBoost identified most important features. As described in previous sections, in some cases including additional variables in the metrics produced improved results while in others it did not. It is well known that many SDHs are statistically and causally intertwined. Without an understanding of possible societal and health causal pathways, purely statistical results cannot be used to improve needs assessments.

## 5 Conclusions

We undertook a validation study of our COVID-19 informed community needs assessment metrics by identifying a proxy measure for the outcome of interest and exploring the predictive power of the underlying variables and other data included in the CVI dataset. This study indicated where some variables needed to be weighted more highly in their respective metrics (‘Covid Cases’ for the Severity Metric, ‘% Self-Employed and ‘Unemployment Rate’ for the Economic Harm metric), identified information gaps in the current metrics (‘% Uninsured’ and ‘% Enrolled in Free and Reduced Lunch’ for Severity, ‘% Severe Housing Cost Burden’ for Economic Harm, ‘Other PCP Rate’ for Mobile Health), and identified variables that should be further studied (‘% Insufficient Sleep’, ‘Social Association Rate’, and ‘Access to Exercise Opportunities’).

We provide a suite of COVID-19 informed community need and risks assessments that can be utilized by non-profits, governments, and community organizations to effectively allocate resources and best support their communities during health crises. This study also has important implications for the construction and validation of quantitative community needs assessment tools. A careful, iterative method is necessary to quantitatively model community needs accurately. With the valuable insights from this study, we will be able to inform further literature review and subsequent iterations of robust metrics.

## Data Availability

Data is all taken from open source databases

## A Initial Variables

**Table 7:**
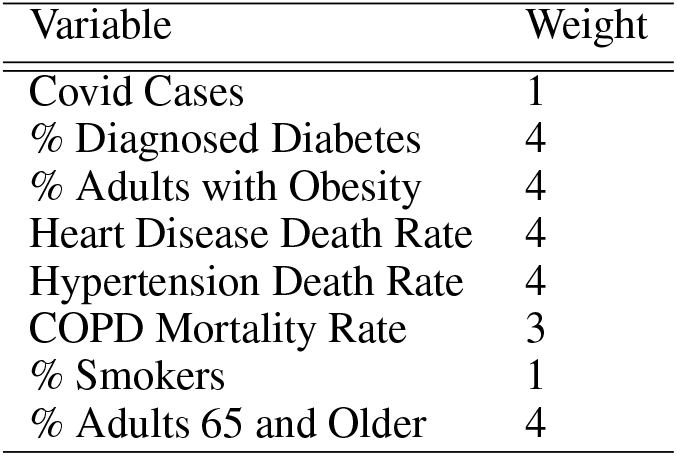
Initial Variables (Severity)

| Variable | Weight |
| --- | --- |
| Covid Cases | 1 |
| % Diagnosed Diabetes | 4 |
| % Adults with Obesity | 4 |
| Heart Disease Death Rate | 4 |
| Hypertension Death Rate | 4 |
| COPD Mortality Rate | 3 |
| % Smokers | 1 |
| % Adults 65 and Older | 4 |

**Table 8:** Initial Variables (Economic Harm)

| Variable | Weight |
| --- | --- |
| % Below Poverty | 1 |
| Median Household Income | 1 |
| % No College Degree | 1 |
| Unemployment Rate | 1 |
| % Not in Labor Force | 1 |
| % Part-time | 1 |
| % Self-Employed | 1 |
| % Jobs in Leisure and Hospitality | 1 |

**Table 9:** Initial Variables (Mobile Health)

| Variable | Weight |
| --- | --- |
| Primary Care Physicians Rate | -3 |
| Number of Hospitals | -3 |
| % Rural | 3 |
| % Households without Car | 2 |
| % Workers Commuting by Public Transit | -2 |
| % Without Health Insurance | 2 |
| % Limited English Proficiency | 2 |
| % Non-white | 1 |
| % Veterans in Civilian Adult Population | 1 |
| % Adults 65 and Older | 2 |
| % People with Disabilities | 2 |
| Opioid Death Rate | 1 |
| % Fair or Poor Health | 1 |

